# When to return genomic newborn screening results: health care professional perspectives

**DOI:** 10.64898/2026.09.02.26362059

**Authors:** Anna C F Lewis, Adam H Buchanan, Aaron J Goldenberg, Bartha M Knoppers, Amy L McGuire, Robert C Green, Ingrid A Holm

## Abstract

**Background and Objectives:** Dozens of projects around the world are sequencing the genomes of healthy babies near birth. Genomic information becomes actionable across infancy, childhood and adulthood, making timing of return a design choice for programs. We report the first study of health care professionals’ views on timing.

**Methods:** We conducted a qualitative interview study of US-based clinical geneticists, genetic counselors, laboratory personnel, pediatric primary care clinicians and genomic screening implementers. Participants responded to three strategies: staged throughout childhood when information becomes actionable, all at birth with adult-actionable results deferred, and all at birth. Transcripts were analyzed using framework analysis.

**Results:** We interviewed 52 individuals; 39 were asked directly about timing. Giving parents a choice, raised by participants rather than presented, was the most endorsed position (18), ahead of staging across childhood (12). Many viewed staging as preferable in theory, but feasibility concerns weighed against it, including that “actionability” was not a robust enough concept. A further concern was that parents would not grasp the distinction between data generated and data examined, on which staging depends. Where information is staged, participants saw a role for adolescent assent; where it is not, disclosure to the developing child becomes important, and participants identified a lack of support for parents.

**Conclusions:** Tying the return of information to the age of actionability is intuitive but hard to operationalize. Parental choice was the most endorsed position but will only be viable with decision support and guidance for disclosure to children.

## Introduction

Sequencing the genomes of healthy babies can enable the identification of many more children that could benefit from early preventative action than standard newborn screening. Newborn sequencing is available through clinical offerings, and over 30 research programs and feasibility studies are currently recruiting babies around the world, including studies in England and New York each aiming to sequence 100,000 newborns.^1–3^ The genome contains more information than is actionable in infancy.^4^ Many genes are associated with conditions for which prevention or surveillance is not required until later in childhood, for example polyposis surveillance from age 10–12 in familial adenomatous polyposis (*APC*),^5^ or only in adulthood, for example surveillance for *BRCA* carriers at age 25.^6^

This leaves open a question of timing: should all clinically actionable information be returned at birth, or staged as it becomes actionable? Existing programs vary: some return all actionable information near birth,^7^ others only information actionable within the first year^8^ or the first five years,^9^ and some are considering whether the data should be re-queried later.^9^ We contrast three strategies (Figure 1): return all genomic results at birth; return childhood-actionable results at birth but defer adult-actionable results to 18, with the individual’s consent; or stage results throughout childhood as they become actionable. These strategies differ in their balance of benefits and burdens, autonomy, feasibility, and privacy. The health care professionals who would deliver and communicate staged information are well positioned to weigh them, but their views have not been studied. We report their views on these strategies, experts’ perceptions of the roles parents and adolescents should play, and how and when children should be informed. These are decisions that will increasingly fall to pediatric clinicians as genomic newborn sequencing scales.

**Figure 1.**
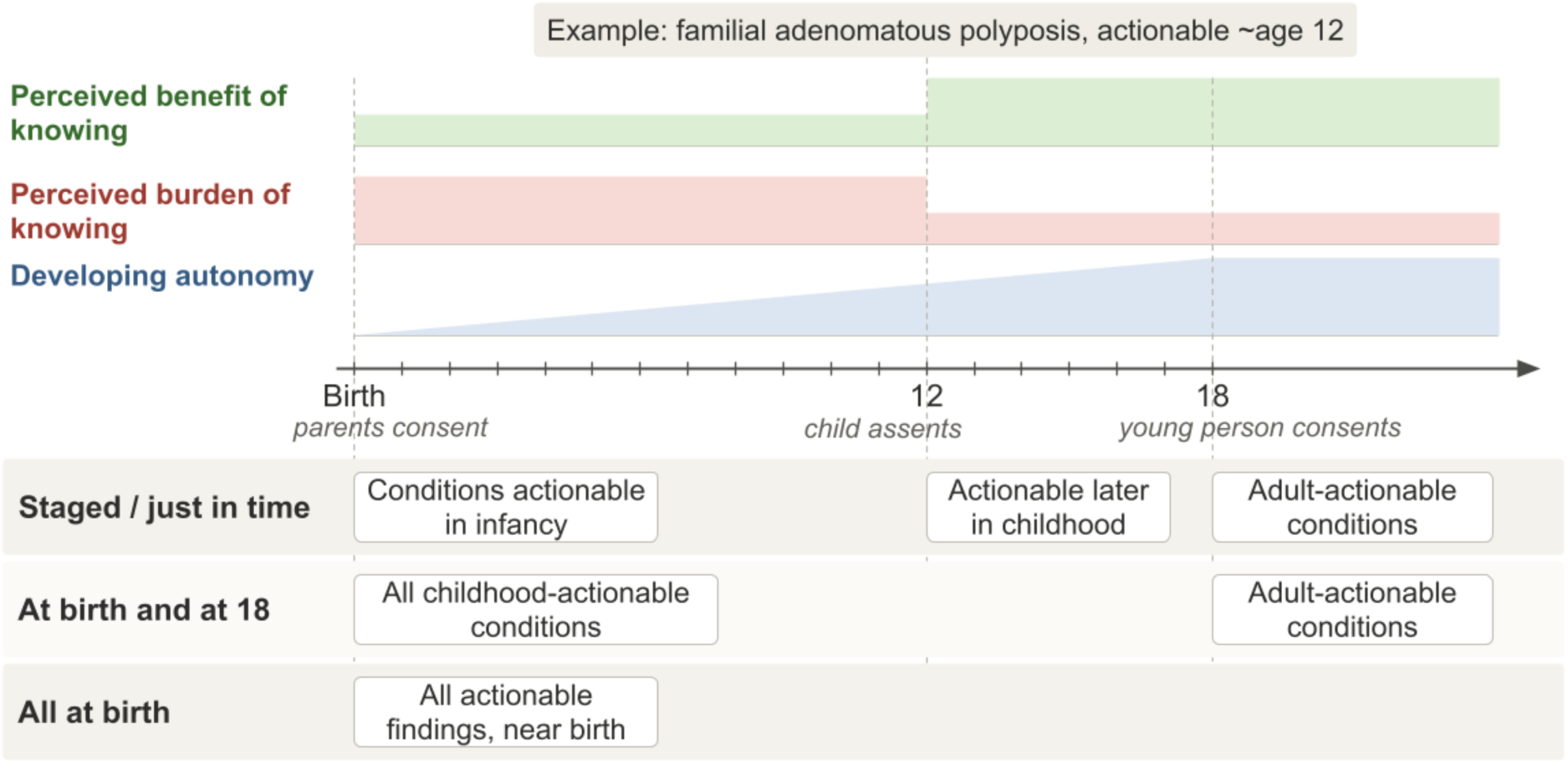
Three strategies for returning childhood genomic findings: staged as conditions become actionable; at-birth-and-at-18 (adult-actionable findings deferred to 18); and all-at-birth. For an example condition (familial adenomatous polyposis), perceived benefit steps up and burden steps down at the age of actionability; autonomy rises throughout childhood.

## Methods

We conducted a semi-structured qualitative interview study with US-based health care professionals: clinical geneticists, genetic counselors, laboratory personnel, pediatric primary care clinicians (PCCs), and genomic screening program implementers. Clinicians were recruited through the recruitment sites of BabySeq,^10^ laboratory personnel through directors of the top ClinVar-submitting laboratories,^11^ and implementers through the leadership of major US genomic screening programs identified from a published review.^12^ Interviews were conducted via videoconference by AL between August 2024 and February 2025.

Full methodological detail and a Consolidated Criteria for Reporting Qualitative Research (COREQ) checklist are provided in Supplementary Methods and Supplementary File 3. The guide (Supplementary File 1) covered experiences of receiving not-yet-actionable information, obligations over time, the clinical value of withholding later-childhood-onset information, expectations of parents and assenting adolescents, and the distinction between generating and querying genomic data; it was used flexibly and tailored to each participant. The three strategies (Figure 1) were not scripted; they were introduced verbally and participants asked to evaluate their feasibility and implications (Supplementary Methods).

Participant age, gender, race and/or ethnicity (per 2024 federal standards^13^), and years since terminal degree were self-reported via a post-interview survey and are reported only to characterize the sample; race and ethnicity are social constructs, not biological variables, and were not used analytically.

We used framework analysis^14^ combining deductive codes from the three-strategy framework (Figure 1) and the guide with inductive codes from responses, conducted in ATLAS.ti. Transcripts were coded by AL, with four co-coded by IH (Supplementary Methods; codebook, Supplementary File 4). Codes on timing of return are reported here; broader lifelong genomic medicine themes and further methodological detail are reported elsewhere.^15^ The Mass General Brigham institutional review board determined the study exempt and did not require documented consent; having reviewed a study fact sheet (Supplementary File 2), participants confirmed before each interview that they wished to proceed. Generative AI (Claude, Anthropic) was used to refine written text; it was not used for data collection, coding, or analysis.

## Results

We interviewed 52 individuals. No substantively new themes emerged in the final interviews within each participant type. Demographic characteristics are summarized in Table 1. Six themes related to whether information should be staged, returned in full at birth, or restricted to childhood-actionable results, with considerations for and against staging distributed unevenly across them (Figure 2; quotes in Supplementary Tables 1 and 2). Participants raised the possibility of giving parents a choice (Supplementary Table 3), endorsed strategies (Table 2), and voiced marked uncertainty.

**Figure 2.**
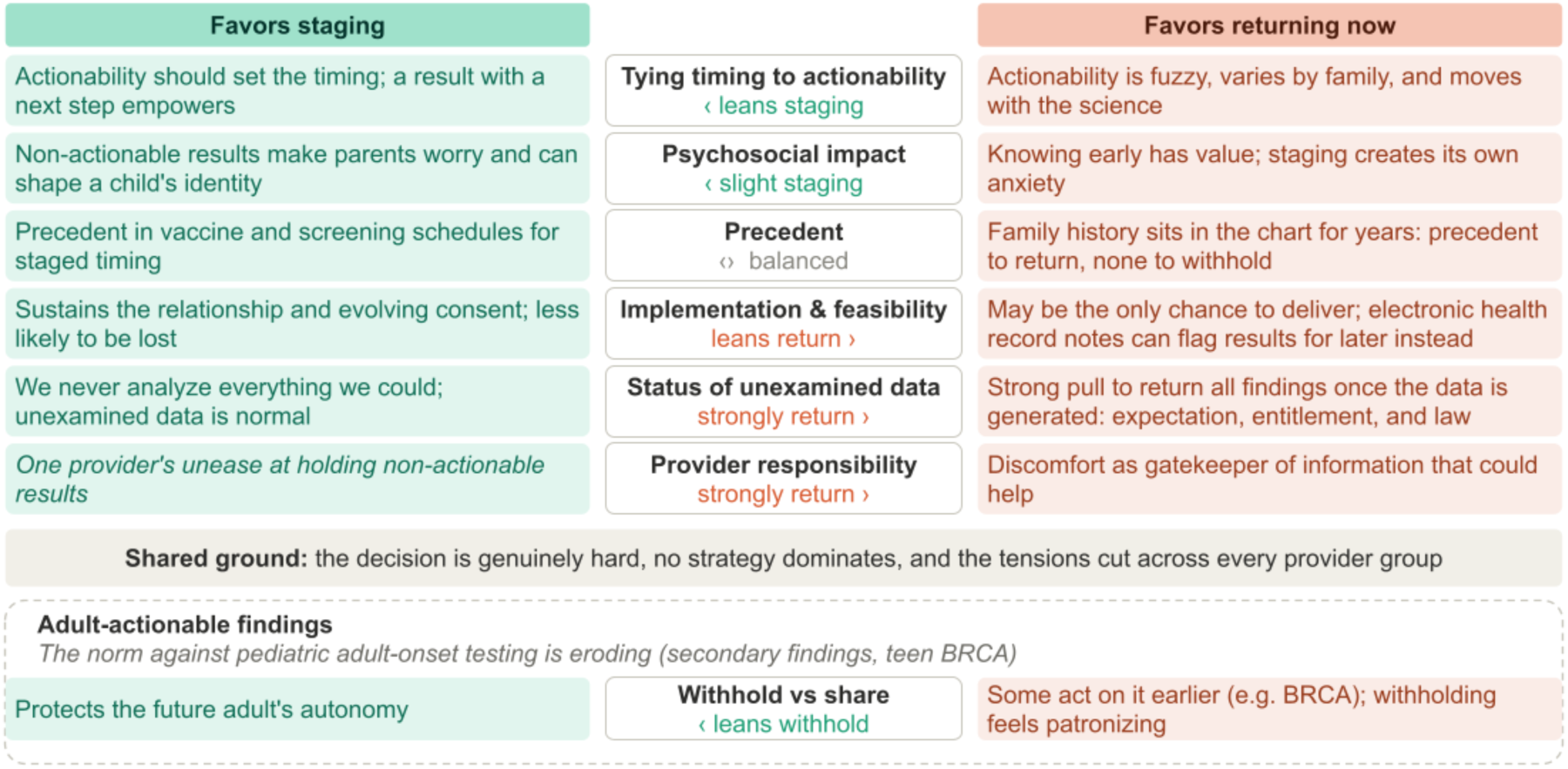
Stakeholder perspectives on staging genomic information over childhood. Six themes drew considerations favoring staged return (left) or near-birth return (right); chevrons mark the more heavily emphasized side. Lean reflects relative prominence, not a count. Convergent views and the adult-onset tension appear below.

**Table 1:** Participant Characteristics. The same interview cohort is described in a companion study.^15^ Demographic data were missing for 4 participants (3 pediatric primary care clinicians and 1 adult genetic counselor) and were excluded from the counts shown. Race and/or ethnicity was self-reported using the combined question and minimum reporting categories of the 2024 revision to OMB Statistical Policy Directive No. 15; respondents could select more than one category.

| Participant group | n | Gender<br>(Female /<br>Male / Prefer<br>not to answer) | Age, mean<br>(range) | Race and/or ethnicity | Years since<br>terminal/highest<br>degree |
| --- | --- | --- | --- | --- | --- |
| Clinical Geneticists | 6 | 2 / 3 / 1 | 50 (39–61) | White: 5<br>Asian: 1 | 11–15 years: 3<br>21–25 years: 1<br>>25 years: 2 |
| Genetic Counselors<br>(adult n=7,<br>pediatric n=7,<br>Maternal fetal<br>medicine n=2) | 16 | 13 / 2 / 0 | 37 (27–56) | White: 10<br>Asian: 3<br>Asian, White: 1<br>Hispanic or Latino: 1 | 0–5 years: 7<br>6–10 years: 2<br>11–15 years: 2<br>16–20 years: 2<br>21–25 years: 1<br>>25 years: 1 |
| Laboratory<br>Personnel | 8 | 1 / 7 / 0 | 52 (40–64) | White: 6<br>Asian: 2 | 6–10 years: 1<br>11–15 years: 1<br>16–20 years: 4<br>>25 years: 2 |
| Primary Care<br>Clinicians (PCCs) | 10 | 4 / 3 / 0 | 48 (28–61) | White: 5<br>Black or African<br>American: 1<br>Hispanic or Latino: 1 | 0–5 years: 4<br>21–25 years: 1<br>>25 years: 2 |
| Genomic screening<br>implementers | 12 | 8 / 4 / 0 | 46 (33–69) | White: 10<br>Black or African<br>American: 1<br>Middle Eastern or<br>North African: 1 | 0–5 years: 2<br>6–10 years: 1<br>11–15 years: 3<br>16–20 years: 1<br>21–25 years: 4<br>>25 years: 1 |
| **Total** | **52** | **28 / 19 / 1** | **45 (27–69)** | **White: 36**<br>**Asian: 6**<br>**Black or African**<br>**American: 2**<br>**Hispanic or Latino: 2**<br>**Asian, White: 1**<br>**Middle Eastern or**<br>**North African: 1** | **0–5 years: 13**<br>**6–10 years: 4**<br>**11–15 years: 9**<br>**16–20 years: 7**<br>**21–25 years: 7**<br>**>25 years: 8** |

**Table 2.**
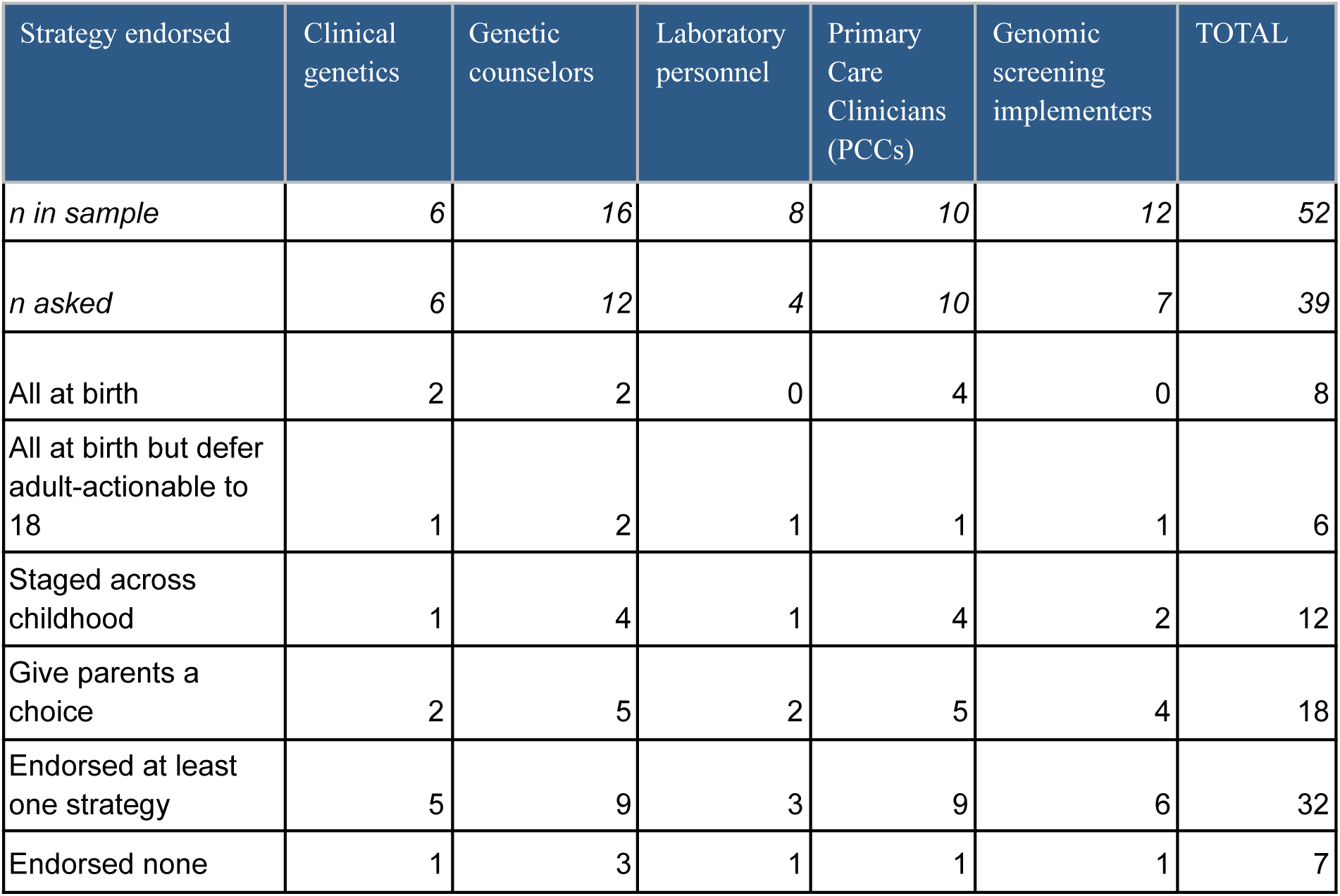
Endorsements for timing strategy, by professional group. Alongside the presented strategies (all at birth; all at birth but defer adult-actionable; staged across childhood), the table includes giving parents a choice, an option raised during interviews rather than presented; because it was not systematically asked about, this count is a lower bound and is not directly comparable to the presented strategies. Cells show numbers of participants. Of 52 participants, 39 were asked their views: 32 endorsed at least one strategy shown here.

| Strategy endorsed | Clinical genetics | Genetic counselors | Laboratory personnel | Primary Care Clinicians (PCCs) | Genomic screening implementers | TOTAL |
| --- | --- | --- | --- | --- | --- | --- |
| *n in sample* | 6 | 16 | 8 | 10 | 12 | 52 |
| *n asked* | 6 | 12 | 4 | 10 | 7 | 39 |
| All at birth | 2 | 2 | 0 | 4 | 0 | 8 |
| All at birth but defer adult-actionable to 18 | 1 | 2 | 1 | 1 | 1 | 6 |
| Staged across childhood | 1 | 4 | 1 | 4 | 2 | 12 |
| Give parents a choice | 2 | 5 | 2 | 5 | 4 | 18 |
| Endorsed at least one strategy | 5 | 9 | 3 | 9 | 6 | 32 |
| Endorsed none | 1 | 3 | 1 | 1 | 1 | 7 |

### Whether to stage information

#### Actionability was the shared criterion but resisted definition

Actionability was the criterion participants across professional groups reasoned with, though they disagreed about what it implied in practice. Two PCCs analogized to not ordering bloodwork unless the result changes management. But actionability was hard to define and operationalize: it depends on circumstances such as family history, resists a universal schedule, and shifts over time (“We don’t know what’s going to be relevant in 20 years from now”; P18, clinical geneticist). One genetic counselor feared everything would default to return near birth because thresholds are so hard to set. PCCs, who already document not-yet-actionable information, considered documenting a result to revisit at a given age itself a form of action.

#### Psychological harm was the leading argument for staging

A widely endorsed argument for staging information was the psychosocial harm of non-actionable information: that parents will worry (the most cited concern) and treat their child differently; that the child may worry; that early disclosure disrupts time with the baby; and that identity can form around a result, *“you’ve gotta let kids be kids”* (P28, pediatric genetic counselor). The counterpoint was the personal utility of more time to prepare, with experiences shared of parents’ relief at knowing and of patients harmed by not knowing. Staging itself may carry costs, replacing a single disclosure with recurring distress at each new result. A few noted the sparse evidence that these psychological harms materialize.

#### Precedents cut both ways

The most cited model for a staging schedule was age-based screening recommendations (eg, the vaccine schedule); others cited precedent for managing not-yet-actionable information, for reanalysis of old genetic data, and for waiting until a result is actionable before testing family members. One implementer countered that medicine has no precedent for holding information back. The norm against returning adult-actionable information to children’s parents was described as shifting and ambiguous: *BRCA* testing is being ordered on teenagers, and secondary findings from sequencing symptomatic newborns already include adult-actionable results. PCCs cited precedent for keeping some information about adolescents confidential from parents.

#### Feasibility concerns dominated the case against staging

The leading feasibility argument against staging was that initial disclosure may be the only opportunity to deliver life-altering information. Participants described losing patients to follow-up and how devastating it would be to miss someone; a childhood finding often identifies an at-risk parent, so delay withholds information from an adult who could act now. This “last chance” framing was contested, given the rising prevalence of genetic testing. The most cited practical obstacle was that people move: “You may move 6 more times… you can’t find a person. Don’t even have the same name.” (P31, PCC); others were that health care systems may not persist over these timescales, and that staging would be confusing to set up and explain. Arguments for staging included that results get buried in the electronic health record (EHR), and that ongoing contact enables evolving consent. PCCs were less concerned about burial in the EHR, detailing how they capture not-yet-actionable information, and suggested EHR-based reminders could prompt action at the right time.

#### Many expected parents not to distinguish data held from data examined

Staging relies on the distinction between data generated and data examined (and disclosed). Considerations here ran overwhelmingly against staging. Participants expected patients, and some providers, to treat unexamined data as nonetheless existing: one called patients’ sense that “they have your DNA” a realistic understanding rather than a misconception (P13, laboratory); another warned of the reaction *“you knew, or you could have known”* (P30, genomic screening program implementer). Some disputed the premise, holding that unanalyzed sequence is no different in kind from blood drawn but not tested: *“you have blood that hasn’t been analyzed, too”* (P39, adult genetic counselor). On this view the distinction could be communicated without difficulty, particularly with pretest counseling. But participants also noted patients could obtain the information via direct-to-consumer testing regardless. Laboratory personnel were vocal about parental rights to know. Separately, a PCC and an implementer raised legal dimensions of access, one holding that once you have the data you cannot not return it. Whether the distinction holds in practice turns on masking, withholding some information from analysis. Laboratory personnel described variable practices, some finding it straightforward and others difficult to do reliably, with one noting they could unmask at will; adult genetic counselors treated laboratory practice as not their concern. For adult-actionable conditions, the dominant argument for withholding was protecting the future adult’s right not to know; the counter was that denying parents access to their child’s DNA-derived information is patronizing.

#### No strategy left providers comfortable

Provider concerns about their role split by profession, though none were comfortable. Genetics professionals voiced “angst” at holding an unreleased actionable finding (clinical geneticists, genetic counselors and implementers). PCCs’ concerns ran the other way: one described anxiety at receiving information that was not yet actionable, worrying about what was being missed by not acting now. A PCC also expressed reluctance to serve as the long-term data steward, seeing this as an unwelcome extension of their clinical role.

#### Many proposed giving parents a choice

Rather than selecting among the strategies, participants across every professional group suggested letting families decide for themselves, noting divergent parent preferences for knowing. *“Some families… ‘I want to know about the* BRCA2 *in my neonate.’ And some… ‘I really don’t want to know’”* (P6, clinical geneticist). Some also described families who, knowing themselves, would prefer not to receive information before it was relevant. Choice was seen as consistent with the many other values-based decisions offered to parents, as improving adjustment (people adjust better to information they chose to receive), as reducing the anxiety that lack of choice can create, and as fairer than having someone else decide. One participant suggested a default option be presented. Several participants who favored choice also named its costs: that choice itself could generate anxiety, that decision fatigue is real, and that offering a choice burdens whoever implements it.

#### No strategy commanded a majority

No strategy was endorsed by a majority of the 39 participants asked, and seven endorsed none of the four options (Table 2). Twelve endorsed more than one, almost always pairing parental choice with a preferred default. Parental choice was the most widely endorsed position overall and in every professional group, tying with all-at-birth among clinical geneticists; staging was next, all-at-birth was concentrated among PCCs, and all-at-birth except adult-actionable was least favored. Two participants endorsed an option not presented: sequencing targeted panels at successive ages rather than storing a whole genome.

#### Many found the choice genuinely difficult

A few argued the question both ways, finding the ethics genuinely unsettled and unsure where the genetic-exceptionalism arguments land. A few made the point that the whole world will change on these timescales. As one participant put it, *“I sometimes worry that there are only wrong answers”* (P18, clinical geneticist).

### Implications for the developing child

Unresolved questions about the developing child’s involvement remain, differing by strategy.

#### The role of adolescents

Participants proposed that, if information were staged, the child should assent to receiving findings at the appropriate stage. Adolescent-parent conflict over genetic testing was described as uncommon, arising when parents wanted testing and the adolescent did not. Participants cited legal precedent for an evolving role, including medical record access at age 13 and minor consent laws for some confidential services. Several argued adolescents should be able to opt into adult-onset testing. Others cautioned that parental authority would remain decisive, describing adolescent assent as potentially “window dressing.”

#### Disclosure to children

PCCs and pediatric genetic counselors agreed that parents bear the responsibility for disclosure of genetic results to their children. *“We say to the parents… when you feel as a parent that your child is ready to hear this… that’s gonna be a new parenting moment for you”* (P3, pediatric genetic counselor). But there was also recognition that there was nothing to support them, and no clear guidelines: *“I’m not aware of guidelines, really, about how to disclose or when to disclose*” (P52, PCC). Some mentioned that earlier disclosure may help children integrate genetic information into their developing identity, and that physicians can prompt parents to have that conversation. One PCC questioned whether the status quo is ethical at all; a few flagged what should happen if the child declines to be told.

## Discussion

Our participants broadly accepted the principle of tying information sharing to actionability, but in practice it is unstable: it depends on personal circumstances, shifts over time, and resists clear age thresholds. Prior work shows the lay public interprets “actionable” far more broadly than geneticists intend;^16^ our data show the same ambiguity among professionals.

The most frequent concern favoring staging, that parents would worry and treat their child differently, maps onto the “patients-in-waiting” phenomenon,^17^ and the vulnerable-child-syndrome literature, including empirical data that newborn screening carrier results for cystic fibrosis and sickle cell increased parental perceptions of child vulnerability.^18^ As some participants noted, these concerns may be overestimated: the BabySeq Project found no undue psychosocial distress from returning newborn genomic results,^19^ though such trials enroll self-selected families and the evidence base remains thin.

Although tying return to actionability was broadly endorsed as a principle, 12 of the 39 participants endorsed staging. This gap suggests that professionals accept the logic of staging more readily than they trust its execution. PCCs and implementers anticipated the recontact burden; geneticists, counselors and implementers felt the moral weight of holding actionable information they could not disclose.

These tensions are particularly sharp for conditions that become actionable in adulthood, where the question is no longer when to return information but whether to return it to a child’s parents at all, rather than to the individual at 18. Here, withholding has a principled basis absent elsewhere. The 2013 American Academy of Pediatrics/American College of Medical Genetics and Genomics (ACMG) statement,^20^ and the American Society of Human Genetics 2015 position statement,^21^ urge deferral of genetic testing unless interventions in childhood reduce morbidity or mortality, while allowing exceptions such as families for whom diagnostic uncertainty poses significant psychosocial burden. That precedent was set for testing children of known carriers, and applies less cleanly to screening families not known to be at risk.^22,23^ Reflecting this, the ACMG secondary-findings policy already recommends the return of some adult-actionable results identified in children.^24,25^ This reflects a turn in the philosophical literature away from a child’s right to an open future^26^ toward a child’s best interests frame.^27^ Our participants’ views reflected this shift: while protecting autonomy was still prominent, many felt restricting parental access was paternalistic.

Allowing parents to decide whether information is staged was the most-endorsed position, despite not being among the three strategies presented. Almost all participants endorsing more than one strategy paired parental choice with a substantive default, suggesting choice was frequently held alongside a preferred schedule rather than instead of one. This recognition of staging as a preference-sensitive, family-level decision reflects the broader shift toward shared decision-making in pediatrics.^28^

The underlying existence of the data may be a faultline for public trust: participants described patients as unlikely to distinguish “we have your genome but only examined part of it” from “we know everything about your genome,” and some regarded the distinction as publicly indefensible. Consistent with this, patients undergoing clinical exome sequencing have contested laboratories’ decisions to withhold information deemed non-actionable, asserting a right to this information rooted in it being theirs.^29^ Parents’ understanding of what genomic sequencing actually examines is known to be incomplete.^30^

A proposed alternative that sidesteps this problem is age-based genomic screening, generating at each successive well-child visit only the data to be examined then, now being piloted in pediatric primary care.^31,32^ In the companion analysis, this narrower approach ran counter to the prevailing view that broad, reusable genomic data is inevitable, with the panel-versus-genome distinction already dissolving as virtual panels increasingly run on exome and genome backbones.^15^

That parents and providers lack support for disclosing results to children (particularly relevant if information is not staged) reflects evidence that parents feel ill prepared for these discussions,^33^ and a recent scoping review identified gaps in genetic health providers’ preparedness.^34^ Developmental guides around specific syndromes for clinicians and parents do exist,^35^ though these presuppose a known familial condition, a symptomatic proband, or an ongoing genetics relationship.

This study is limited to provider views; parents and children are not represented. It is limited by being US-based and recruited largely through BabySeq sites, so views may reflect a fragmented, mobile, multi-payer system; feasibility objections may not transfer to single-payer settings. The framing was hypothetical. Because strategies were introduced responsively rather than as a fixed instrument, and because giving parents a choice was raised by participants rather than presented, the counts in Table 2 describe the distribution of views among those asked rather than prevalence in the sample. Most coding was conducted by a single researcher, which may have traded analytic breadth for interpretive consistency.

## Conclusion

As more healthy babies are sequenced, every program must decide what happens to information that becomes actionable after infancy. One possible future stages this information across childhood, sharing it with parents as it becomes clinically actionable. The obstacles to this are largely operational, contingent on health system features such as continuity of care and record persistence, and therefore potentially tractable. Returning results near birth that become actionable only later runs against norms in medical genetics, yet there are compelling reasons not to treat age of actionability as the cutoff, letting other screening criteria such as the overall balance of benefits to harms govern instead. If parental choice is adopted, decision-support scaffolding (defaults, values-clarification) and disclosure guidance will be needed, so the choice doesn’t collapse into decision fatigue or an unsupported burden. Irrespective, pediatric and laboratory professional guidance are urgently needed to clarify responsibilities and protect the “actionable” health interests of all newborns in genomic newborn screening programs.

## Acknowledgements

We thank all participants for their time, and Ali MacLeod, Mai Ly Burke, Julia Mizzi, Jazmine Harry, Yuka Kato, Layla Horwitz, and Lily Hamilton for their careful work cleaning transcripts. During preparation of this manuscript, the corresponding author used Claude (Anthropic) to refine written text; generative AI was not used for data collection, coding, or analysis, and the authors take full responsibility for all content.

## Abbreviations

PCC: primary care clinician
EHR: electronic health record
ACMG: American College of Medical Genetics and Genomics
COREQ: Consolidated Criteria for Reporting Qualitative Research

## Conflict of Interest Disclosure

Dr Green receives compensation for advising Allelica, Mammoth Biosciences, and Genomic Life, and is a co-founder of Genome Medical and Nurture Genomics. The other authors have no conflicts of interest relevant to this article to disclose.

## Funding/Support

This work was supported by the National Human Genome Research Institute (award K99HG012809 to Dr Lewis). The funder had no role in the design or conduct of the study; collection, analysis, or interpretation of the data; or preparation of the manuscript.

## Data sharing statement

Deidentified individual participant data will not be made available. Interview transcripts cannot be shared without risk of re-identification, given the small, specialized professional community; the interview guide, coding framework, and COREQ checklist are provided as supplementary material.

## Contributors’ Statement

Anna Lewis conceptualized the study, acquired funding, conducted all data collection and curation, led the formal analysis, administered the project, created all visualizations, and wrote the original draft. Ingrid Holm contributed to the formal analysis, provided supervision, and conducted validation. Robert Green acquired funding and provided supervision. Adam Buchanan, Aaron Goldenberg, Bartha Knoppers, and Amy McGuire contributed to the methodology. All authors reviewed and edited the manuscript, approved the final manuscript as submitted, and agree to be accountable for all aspects of the work.

## Supplementary information

**Supplementary Methods.** Extended methodological detail: research team and reflexivity, sampling and recruitment, data collection, analysis, and reporting.

**Supplementary Table 1-3.**

**Supplementary File 1. Interview guide.** The semi-structured guide used across all five professional groups, developed from the literature and the research team’s expertise and tailored to each participant.

**Supplementary File 2. Study fact sheet.** The information sheet describing the study and its purpose, provided to all prospective participants before they agreed to be interviewed.

**Supplementary File 3. COREQ checklist.** Completed Consolidated Criteria for Reporting Qualitative Research checklist, with the location of each item in the manuscript.

**Supplementary File 4. Codebook, with counts**. The complete coding framework, with code frequencies by stakeholder group.

### Supplementary Methods

#### Research team and reflexivity

All interviews were conducted by AL (DPhil, she/her), a researcher working at the intersection of genomics and ethics, and were part of her research program. AL was a partial insider to the professional community studied, which may have facilitated candor but may also have led some participants to foreground ethical considerations. A small number of participants were known to AL before the study; all were assured that their responses would be kept confidential. IH (MD) is a clinical geneticist and researcher whose expertise provided an additional analytical perspective and helped surface assumptions that might otherwise have gone unexamined. Both AL and IH are experienced qualitative researchers.

#### Sampling and recruitment

We used purposive sampling, supplemented by snowball sampling in which interviewees were asked to suggest others within their institutions who might hold differing perspectives or relevant expertise. Five professional groups were recruited to enable triangulation: clinical geneticists, genetic counselors (pediatric, adult, and maternal-fetal medicine), laboratory personnel (predominantly laboratory directors), pediatric primary care clinicians, and genomic screening program implementers (individuals with operational leadership roles integrating genomic screening into health systems, some of whom were clinicians and some primarily administrators). Clinical participants were recruited from the four BabySeq Project sites: Boston Children’s Hospital; the University of Alabama Medical Center, Birmingham; Corewell Health, Detroit; and Children’s Hospital of Philadelphia, including affiliated academic and community clinics. Genetic counselors were also invited from the University of Pennsylvania Health System, a sister institution to Children’s Hospital of Philadelphia. Invitations were distributed by email, some individualized and some via institutional listservs; response rates could not be calculated for listserv-recruited groups because the number of recipients was not ascertained. Laboratory personnel were identified from the top submitters of variants to the ClinVar database and invited by email to their directors (7 of 14 participated; one further interviewee was recruited via snowball sampling). Genomic screening implementers were identified from a narrative review of US population genomic screening programs, supplemented by programs known to the research team; of 18 individuals invited, 12 participated. All prospective participants received a fact sheet describing the study and its purpose before agreeing to participate (Supplementary File 2).

#### Data collection

Semi-structured interviews were conducted by AL between August 2024 and February 2025 over Zoom videoconferencing, and were approximately 60 minutes long. Only the interviewer and participant were present. Interviews were recorded; auto-generated transcripts were manually cleaned and de-identified to protect confidentiality. Field notes were taken immediately after each interview. Transcripts were not returned to participants, and no repeat interviews were conducted. A short demographic survey was emailed to participants after the interview, and participants received a $100 honorarium. The interview guide (Supplementary File 1) was developed from the literature and the research team’s expertise and was designed to be tailored to each participant. Several lines of questioning were developed during interviewing rather than scripted: the three candidate timing strategies (Figure 1) were introduced verbally, usually alongside the question on using the genome as a resource over the lifespan; probes on returning adult-actionable results to parents, and on disclosure to children, developed as these issues recurred. Strategies a participant had already addressed were not re-presented, and where an interview centered on curation or implementation the strategies were not introduced. Some guide components were common to all groups; others were specific to clinical versus operational roles.

#### Analysis

Analysis drew on framework analysis, combining deductive and inductive elements, with a pragmatist orientation prioritizing findings of direct relevance to policy and practice. Domains were defined a priori from the three-strategy framework and interview guide. Within each domain, de-identified transcripts were coded in ATLAS.ti using in vivo and descriptive codes. Codes were iteratively grouped into themes and subthemes through repeated comparison across codes, transcripts, and stakeholder groups, and the coding framework was revised over multiple rounds; code frequency was tracked by stakeholder group to enable systematic comparison of where perspectives converged and diverged. Transcripts were coded by AL, with four transcripts co-coded by IH to identify themes and refine the codebook. AL maintained analytic memos throughout, and emerging themes were sense-checked through discussion with research colleagues. No substantively new themes emerged in the final interviews within each participant type. Participant checking of findings was not conducted; given the multi-stakeholder design and the analytic focus on cross-group comparison, we prioritized peer debriefing and co-coding over individual participant validation. The complete coding framework, with code frequencies by stakeholder group, is provided in Supplementary File 4. Codes relating to lifelong genomic medicine broadly, and to population genomic screening, are analyzed separately.

#### Reporting

This study is reported in accordance with the COREQ checklist (Supplementary File 3). Participant quotations are presented throughout the Results and identified by participant number and professional group.

---

**Supplementary Table 1.** Stakeholder Perspectives on Whether Genomic Information Should Be Staged Over Time.

| Theme | For Staging | Against Staging |
| --- | --- | --- |
| Actionability | *“The rule of thumb is you only screen if you can get reliable information, and then that changes your plan of care in some way [...] information that is not directly applicable to the clinical question that you’re trying to answer is not useful information.”* (P47, PCC)<br>*“[Staging] is a really neat idea [...] space it out to when it becomes actionable. I really like being able to give people information that’s empowering [...] because there then is a next step.”* (P28, Genetic Counselor-pediatrics) | *“[Staging is] very logistically challenging, because [...] someone has to make decisions about what age is this information useful.”* (P52, PCC)<br>*“I don’t think the onset ages are as clear as that [...] some of the conditions that we think of as adult onset [...] they can present in childhood.”* (P22, Genomic screening implementer)<br>*“More often than not patients feel the value of this information can go beyond the tangible screening. There’s the mental aspect [...] family planning can be another motivation.”* (P37, Genetic Counselor-adults) |
| Psychological Impact and Uncertainty | *“Parents start to worry should I put them in sports? Should I let them go to summer camp? [...] you’ve gotta let kids be kids.”* (P28, Genetic Counselor-pediatrics)<br>*“If we test a three-year-old and screening doesn’t begin until they’re 10, having seven years of conversations of ‘we’re not doing the MRI now’ [...] [is] difficult for their emotional ability.”* (P14, Genomic screening implementer)<br>*“If you gave them too much info, they’re not going to be able to focus on that baby. They’re gonna worry about that kid every second of the day.”* (P44, PCC) | *“[There is] personal utility in knowing potentially prior to the age of medical actionability [...] they’d rather know versus not know, so the personal utility of putting to bed the uncertainty is really powerful.”* (P35, Genetic Counselor-adults)<br>*“The whole concept of [...] you have to wait till your tenth birthday, and then you get this information, is very stressful [...] is there gonna be a bomb that drops on my eighteenth birthday?”* (P34, Genetic Counselor-pediatrics)<br>*“There are harms of knowing something before it’s actionable. There are psychological harms. There are also psychological harms in not knowing, and sometimes that can bother you more than knowing.”* (P39, Genetic Counselor-adults) |
| Precedents | *“If public policy says no, then there’s not much that can be done about it, and people will just accept it in the same way that they accept that they have to pay for their luggage [...] it’s unpopular initially, and then just becomes part of day-to-day life.”* (P2, Clinical geneticist)<br>*“There are existing precedents for setting ages of actionability [...] it’s like their eight-year-old checkup, by the way, now we’re going to screen for these other things.”* (P14, Genomic screening implementer) | *“If we had a way of predicting that someone was going to get coronary artery disease or fibrotic lung disease, would you wait until the moment when it becomes important to tell them? No, you just tell them. So why is genetics so blankety-blank different?”* (P26, Genomic screening implementer)<br>*“For [...] the newborn screen, if it’s just a carrier [...] it really won’t affect our patient. But later in life, when they get married and decide to have kids, it may affect them. We anyway tell them.”* (P49, PCC) |
| Implementation and Feasibility | *“My hope is always that when the child’s ready to have kids of their own, that they’ll meet with a genetic counselor [...] but my guess is probably a lot of these are forgotten.”* (P14, Genomic screening implementer)<br>*“[Staging] allows for more of an ongoing discussion with the family [...] so that they can take more ownership and more like an evolving informed consent process.”* (P14, Genomic screening implementer)<br>*“[Releasing] information at the age that it’s relevant [...] is not only helpful for families, cause they can kind of prioritize where their energy goes, but also for providers, so things don’t get lost.”* (P51, PCC) | *“This is the bite at the apple. You’re not going to get a second chance. They may never come back [...] they need to be told, ‘this is something that when the kid is 40 years old he needs screening for.’”* (P18, Clinical geneticist)<br>*“You may move 6 more times [...] you can’t find a person. Don’t even have the same name.”* (P31, PCC)<br>*“We put it in the problem list [...] should be referred to genetics at age 5 or age 10, or whatever.”* (P49, PCC) |
| Status of Unexamined Data | *“I don’t think people know enough about what [genomic sequencing] can tell you that then they’re like anxiously waiting for the day that they can run that test. It would just be more like, ‘we’re gonna do it. And if we see something, we’ll tell you. ‘Sort of like the newborn screen.”* (P51, PCC) | *“In the legal framework in the United States, parents are the guardians of their children, and to say a laboratory can have data on somebody’s child but the parents can’t have that data just flies in the face of how we think in our society.”* (P13, Laboratory)<br>*“There are families and individuals and parents who want to just know it all. You cannot deny them that information.”* (P21, Laboratory) |
| Provider<br>Responsibility | *“[When asked about receiving non-actionable results:] Why can’t we do something now? What are we missing? [...] I’m hoping I’m not missing anything.”*<br>(P46, PCC) | *“I shouldn’t necessarily be the guardian of their information. I can be a counselor [...] but I don’t want to be the housekeeper of that information.”* (P33, PCC)<br>*“We had this variant that we knew about, and it was sitting there, and we couldn’t do anything with it [...] I do struggle.”* (P11, Genomic screening implementer) |

**Supplementary Table 2.** Stakeholder Perspectives on Withholding Versus Sharing Adult-Onset Genomic Findings.

| Theme | For Withholding Adult-Onset | For Sharing Adult-Onset |
| --- | --- | --- |
| Precedents and Framing | *“We don’t test kids for adult-onset conditions unless there are early intervention implications.” (P36, Genetic Counselor-pediatrics)*<br>*“BRCA is an ethical challenge. We would report it if it came as a secondary finding. What we wouldn’t do is test for it as a primary test [...] a mother or family member is BRCA positive and they want to get their kids tested. And then we say, no.” (P2, Clinical geneticist)*<br>*“About 50% of the time, we get pretty substantial pushback [...] we lean into [...] we want the child to be a child. We don’t want to medicalize the child.” (P14, Genomic screening implementer)* | *“[BRCA testing in teenagers:] they do, mostly in teenagers [...] some families do engage in surgical procedures afterward or fertility preservation [...] kids or young adults do act on them quite often.” (P32, PCC)*<br>*“The answer was always ‘absolutely not. Wait till you’re 18, and then you can decide. ‘ And I always had an issue with that [...] it shouldn’t always be a hard no.” (P25, Genetic Counselor-maternal-fetal medicine)*<br>*“[If I knew a patient carried a BRCA variant] I could help get them into some type of earlier counseling [...] make sure that you get appropriate medical care, make sure you do the appropriate screening.” (P33, PCC)* |
| Autonomy, Rights and Access | *“I’m worried about [...] creating a kind of vulnerable child syndrome [...] what right do the parents have to know about a mutation [...] that the patient will have the opportunity to make decisions about when he or she is an adult?” (P52, PCC)*<br>*“The child should have the autonomy [...] to decide if they want to learn this information in adulthood. Parents often are very interested [...] they want to test their children, and I have to explain it’s important to protect their child’s autonomy.” (P37, Genetic Counselor -adults)*<br>*“It’s probably not appropriate to return a BRCA1 mutation at birth. Like, what are you going to do with that? And then you’ve kind of taken away the patient’s ability to choose whether they want to have that information.” (P24, Genomic screening implementer)*<br>*“[A parent said] ‘my kid always wanted to be a ballerina and I wanted her to have a secure job with good life insurance in case she was at high risk. ‘ And the parent didn’t let that happen.” (P39, Genetic Counselor-adults)* | *“It’s paternalistic and inconsistent with patient rights to data access [...] if you have my data and it’s about me, then [...] who are you?” (P10, Laboratory)*<br>*“[Withholding from parents] feels patronizing [...] ‘we’re only going to give you what you need to know immediately. We’re not giving you the rest’ [...] they’re adults. They can make decisions [...] they have the right to make decisions for their child.” (P52, PCC)*<br>*“I wonder if families or people will find that very paternalistic? [...] to be told there’s nothing at age 2, or at the beginning, and then, at age 5 [...] ‘now we have this information that we chose not to [share]’ [...] I can see a rationale [...] but I also think it’d be very hard for people to understand when you try to implement it.” (P5, Clinical geneticist)* |

**Supplementary Table 3.** Stakeholder Perspectives on Whether Parents Should Be Given a Choice About the Timing of Genomic Information.

| Theme | Illustrative Quote(s) |
| --- | --- |
| *Arguments for giving parents a choice* |  |
| Variable patient preferences for knowing | *“Some families are like, ‘I want to know about the BRCA2 in my neonate.’ And some are very much like, ‘I really don’t want to know about that’ [...] I would be hesitant to have a provider decide that [...] some families just can’t get these things out of their heads [...] Some families might just say, ‘I just know myself, and I’d rather not know this until it’s relevant.’” (P6, Clinical geneticist)*<br>*“Staring into the future can be a little scary for people [...] there are people out there who are gonna say, ‘I don’t want to know any of this’ [...] we’re gonna have some big group of people in the middle, who are interested in some but maybe not all.” (P27, Genomic screening implementer)*<br>*“Some people want to know for family planning or personal reasons, while others don’t want to know at all [...] Some people don’t want to look for problems, while others want all the information they can get. All types of people exist.” (P36, Genetic Counselor-pediatrics)* |
| Standard to put values-based choices to parents | *“I like the idea of not being overly paternalistic about it and giving them the choice [...] there’s a middle ground, which is to say, ‘here’s choices A and B for you. And the reasons a person might choose A is because they view the world this way’ [...] trying to explain to people why one would choose A or B, like what values go into the choice, helps them to frame it.” (P52, PCC)*<br>*“Could you give parents that option upfront? [...] that’s putting it all on the parents versus the child, but [...] we do that all the time. We do take away autonomy with this testing and letting the parents decide about secondary findings.” (P42, Genetic Counselor-pediatrics)* |
| People adjust better to information if given the choice | *“Psychology studies show us that when people have made that choice, they tend to adjust better because they had the power to choose [...] versus being forced to do this large panel on their child, and then feeling blindsided with information that they might have otherwise not chosen.” (P37, Genetic Counselor-adults)* |
| Lack of choice as anxiety-provoking | *“It’s pretty paternalistic to say ‘you just don’t get this information because nobody does’ [...] having the option to know might even make it more palatable [...] versus someone being like, ‘we’re gonna decide when you get it.’ Like, to me, that just automatically makes me anxious.” (P25, Genetic Counselor-maternal-fetal medicine)* |
| Should be the parents' choice | *“It’s not fair to necessarily say, like the hospital system or another person gets to decide that for somebody else. It feels like a decision people should be able to make for their children or for themselves.” (P5, Clinical geneticist)* |
| There could be a default choice | *"I do think there should be a recommended-by-committee schedule of when certain types of information are released. It shouldn't be automated [...] You have to be informed and say, 'we are planning to release this information. Do you want this information?'"* (P3, Genetic Counselor-pediatrics) |
| *Arguments against giving parents a choice* |  |
| Offering choice can create psychological burden | *"Some people, just like being asked that question, knowing there's something they might not know, will make them anxious."* (P5, Clinical geneticist)<br>*"Decision fatigue is very real [...] just giving people a whole lot of options is hard, because people may say, 'I know I'm supposed to be making a decision, but I don't feel like I have all the information I need' [...] it can just become a lot for families."* (P30, Genomic screening implementer) |
| Offering a choice is a lot for a health system | *"Maybe we need to take into account both pieces. So patient choice, patient preference, and then also what's best for the system, and figuring out how we maximize both of those things."* (P12, Genomic screening implementer) |

## Interview guide for “An interview study to help assess the clinical value and feasibility of revealing genetic information at multiple points of the lifespan”

*This Interview guide should be tailored to the specific respondent I am interviewing*.

***For everyone***

**Preamble**

Hello, []. Thank you for agreeing to take part in this interview study. My name is Anna Lewis and I’m a research scientist at Brigham and Women’s Hospital division of genetics. My mentor is Dr Robert Green and he is the PI of this study.

As a reminder, we are conducting a research study. The purpose of this interview is to help assess the value and feasibility of revealing genetic results over the lifespan. There are no right or wrong answers to any questions.

I sent you a fact sheet about this study. In brief, this interview will be recorded and then transcribed and analyzed. All identifying information will be removed and all of the analyzed data will remain anonymous. This interview will last about 60 minutes. For participating in this interview, we will send you a $100 check as a thank you for your time. Your participation in this interview is completely voluntary and if at any point you wish to stop the interview, you are welcome to do so. Additionally, if there is a question you would rather not answer, just let me know.

As a thank you for your time, we will send you a $100 check. Per hospital policy, I need to ask you for your SSN in order to issue the check. I’ll do so at the end of the interview; it will be used solely for this purpose, kept secure, and will not be connected to your responses.

Before we get started, do you have any questions about what I have presented, or any questions about the Fact sheet?

Do you consent to be part of this study? Is it okay if I begin recording now?

**Introduction to the Project:** Frame some of the key pieces of background, as well as the motivations and concerns. Explain why they were selected to be interviewed.

**1. Background Questions:** *This basic set of open ended questions are designed to get the respondent comfortable sharing and telling us about their work.*

- What is the focus of your professional practice?
- What led you to this area?

***For health care providers (Aim 1a)***

**2a. Reactions to receiving screening information.**

First ask for examples of where they receive both genetic and (where appropriate) non-genetic examples. For these examples, ask:

- What makes it useful?
- What makes it a burden?
- How does uncertainty factor in?
- Examples of receiving information before they would initiate a risk mitigation strategy?

- Idea of “patients in waiting”

**3a. How they view their obligations over time to patients and their parents**

- Example of a variant of uncertain significance for a child with a phenotype
- Example of a child with a pathogenic variant but not yet showing signs of having the condition (low penetrance?)

**4a. Their experiences of longitudinal relationships with patients and their parents**

- Any experiences of patient-parental divergence of opinion with regards to genetic knowledge?
- Any observations of the resources families need access to in order for the value of genetic information to be realized?

**5a. Attitudes toward when information should be shared.**

- Are there example genetic conditions where you think it would be optimal to do population screening later in childhood?
- In other words, do you see any clinical value in withholding any later childhood onset information?

**6a. Their expectations of what patients and parents would want**

- What would you anticipate parents would want when it comes to just-in-time information, versus receiving it all at once?
- What about patients themselves, as they reach the age of assent?

***For those with operational roles in generating genomic reports (Aim 1b): not all questions will be relevant to all roles***

**2b. [Lab personnel] Treatment of sequence information after an initial report is generated**

- Current treatment -- does anything happen? For example, reinterpretation of variants?
- Do you envision this changing in the nearterm, and why?

**3b. [Those embedded in health system] Vision for integration of genomic information into health system**

- What is the near term vision for the integration of genomics into the health system?
- [If not in the near term vision] Do you share a medium term vision of using the genome as a resource over the lifespan? Why or why not?

**4b. How they view their obligations to the individuals who they provide testing for**.

- Does the lab have any legal or regulatory responsibility to the individuals you provide reports for, after the report is signed out?

- What about reporting variants that have taken on a new significance?
- Do you think the lab has any *ethical* responsibility in this area?
- Do you think anyone else, perhaps the ordering physician, has any ethical responsibility in this area?

**5b. [Lab personnel] Whether, and how, their lab currently distinguishes in theory and in practice genetic information from the querying of that information.**

When a lab generates a variant file, in some sense there is information that is both “there and not there” -- a simple look up could reveal a pathogenic variant, but it won’t necessarily be looked up.

- Does your lab distinguish in theory or in practice genetic information from the querying of that information?

**6b. Privacy, security and longevity of information.**

- What do you consider necessary, currently to maintain the privacy, security and longevity of sequence information?
- Do you see any of this changing in the near or mid term?

***For everyone***

**7. Their views on using the genome as a resource over the lifespan**

*[The three candidate timing strategies shown in Figure 1 were typically introduced at this point and participants asked to evaluate them. Probes on returning adult-actionable results to parents, and on disclosure to children, were developed during interviewing.]*

- Do you think that this vision, of sequencing once and querying many times, is one that the field should be pursuing?

- Why or why not? Any considerations we haven’t covered? (anticipate a mixture of practical and ethical concerns)
- Any additional practical considerations that we haven’t touched upon?
- If ethical concerns haven’t come up explicitly: Do you have any ethical concerns?
- If equity hasn’t come up: Do you foresee any equity barriers?

**8. Wrap up.**

- Is there anyone else you recommend I should talk to about these kinds of questions? Particularly someone who might have a contrasting view to your own?

- If so, can I send you a recruitment email, for you to forward to this/these individuals, copying me?
- Is there anything else you would like to add?
- Do you have any questions for me?

I am going to stop recording now. Please can I ask you for your SSN? I will store this securely and we will only use it to issue your check. Can I also please confirm what address you would like the check sent to?

Thank you!

## Mass General Brigham

Study Title: An interview study to help assess the clinical value and feasibility of revealing genetic information at multiple points of the lifespan

Principal Investigator: Robert Green, MD, MPH

Fact Sheet

- This is a Mass General Brigham research study. The purpose of this research is to help assess the value and feasibility of revealing genetic results over the lifespan. This research study is funded by the NIH.
- We are asking you to participate because you are a professional with relevant expertise to help the research team address their research questions. About 64 people will participate in total.
- If you are being emailed directly by the study team, your email was obtained from publicly available information, given your professional role in either a genomic testing lab or a program integrating genomic screening into health systems.
- In this study we will use the platform Zoom to conduct a one-on-one virtual interview. This virtual interview will take 60 minutes and will be recorded. We will ask you questions that draw on your professional expertise; there is no preparation needed on your part.
- You will receive a $100 check for your time. We will need to collect your social security number and address to issue the check. This information will be kept secure and will not be used as study data.
- This interview will be transcribed and analyzed by the study team. We may use direct quotes from participants when presenting our findings.
- The video recordings will only be available to the study team. Your de-identified information may be used or shared with other researchers without your additional informed consent.
- The main risk is a potential loss of privacy and/or confidentiality. We have multiple procedures in place to protect privacy and confidentiality. While we cannot guarantee that the identity of the participants will not be revealed in the event of a breach or other unexpected event, we have precautions in place to keep your information secure. Study data will only be accessible to study staff; your identifiers will be separated from your responses and all data will be coded. We will use the secure systems at Mass General Brigham, endorsed by our Information security team, to secure both your study data and the information provided for issuing the check.
- Participation is voluntary and you can stop at any time.

Anna C F Lewis, DPhil, is the main point of contact for this research study. You may contact her through email at. The PI’s contact information is. If you’d like to speak to someone not involved in this research about your rights as a research subject, or any concerns or complaints you may have about the research, contact the MGB Human Research Committee at (857) 282-1900.

Version: 1/24/2024

### COREQ (Consolidated Criteria for Reporting Qualitative Research) Checklist

*When to return genomic newborn screening results: health care professional perspectives. Based on Tong A, Sainsbury P, Craig J. Consolidated criteria for reporting qualitative research (COREQ): a 32-item checklist for interviews and focus groups. Int J Qual Health Care. 2007;19(6):349-357*.

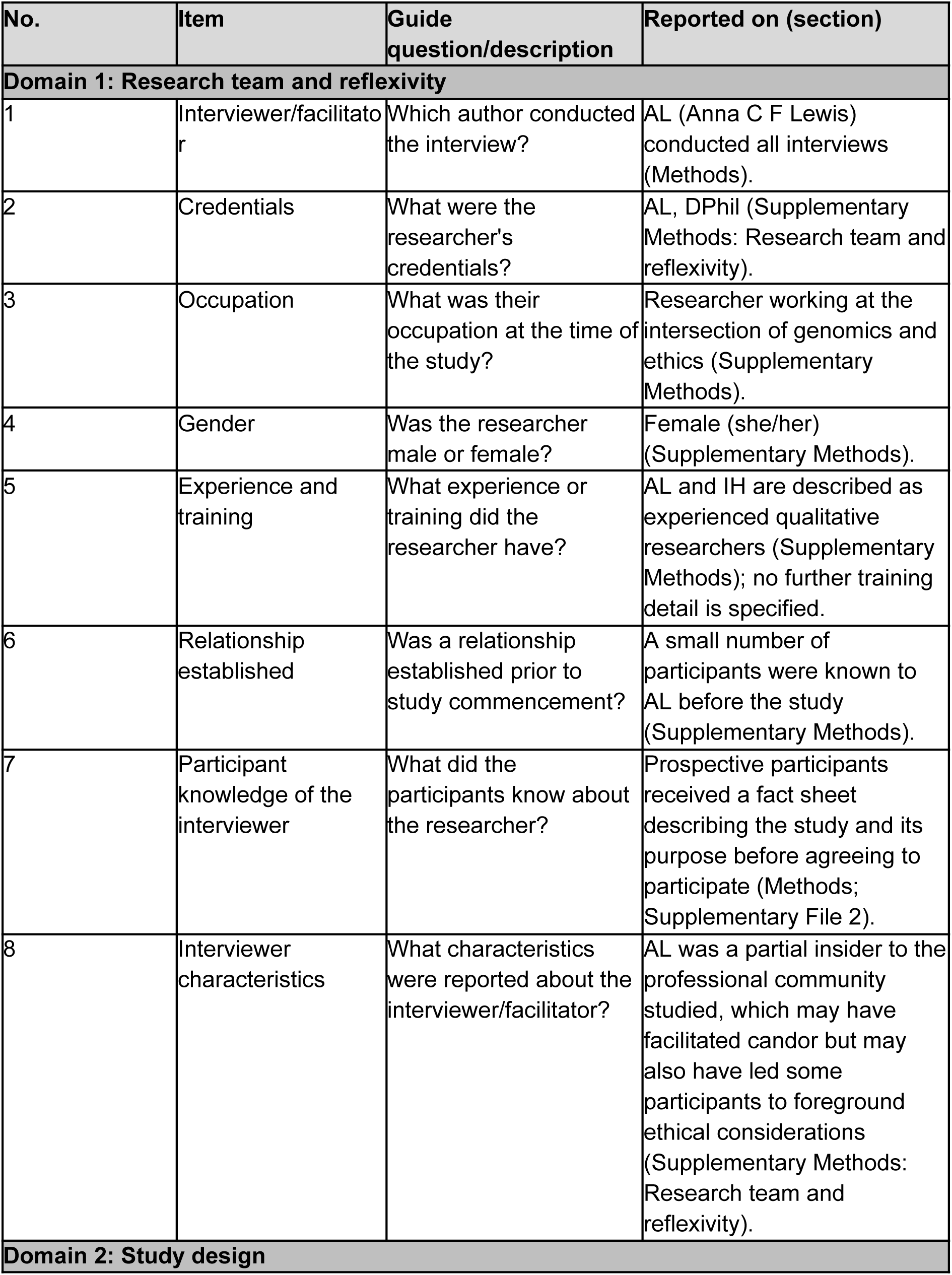
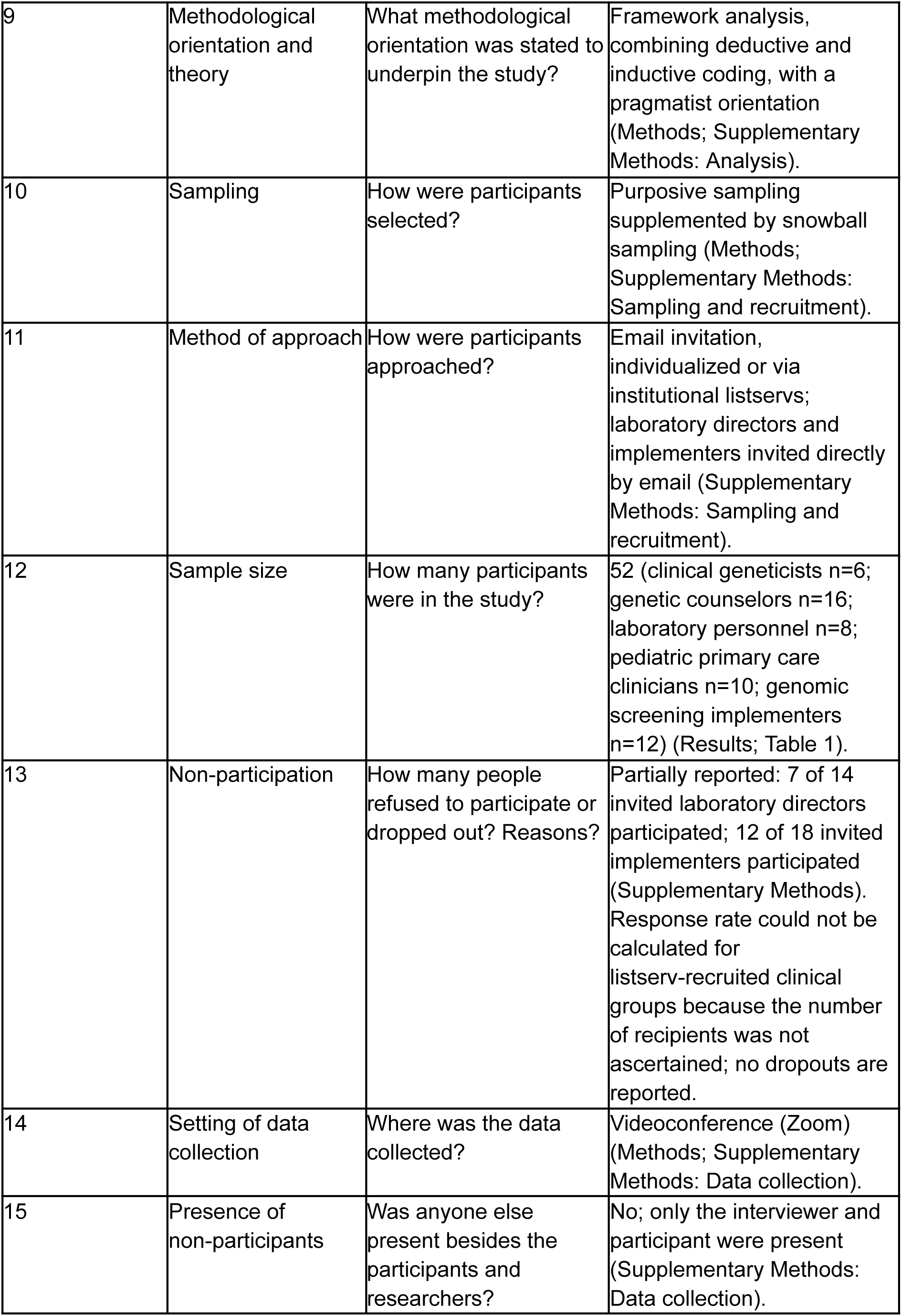
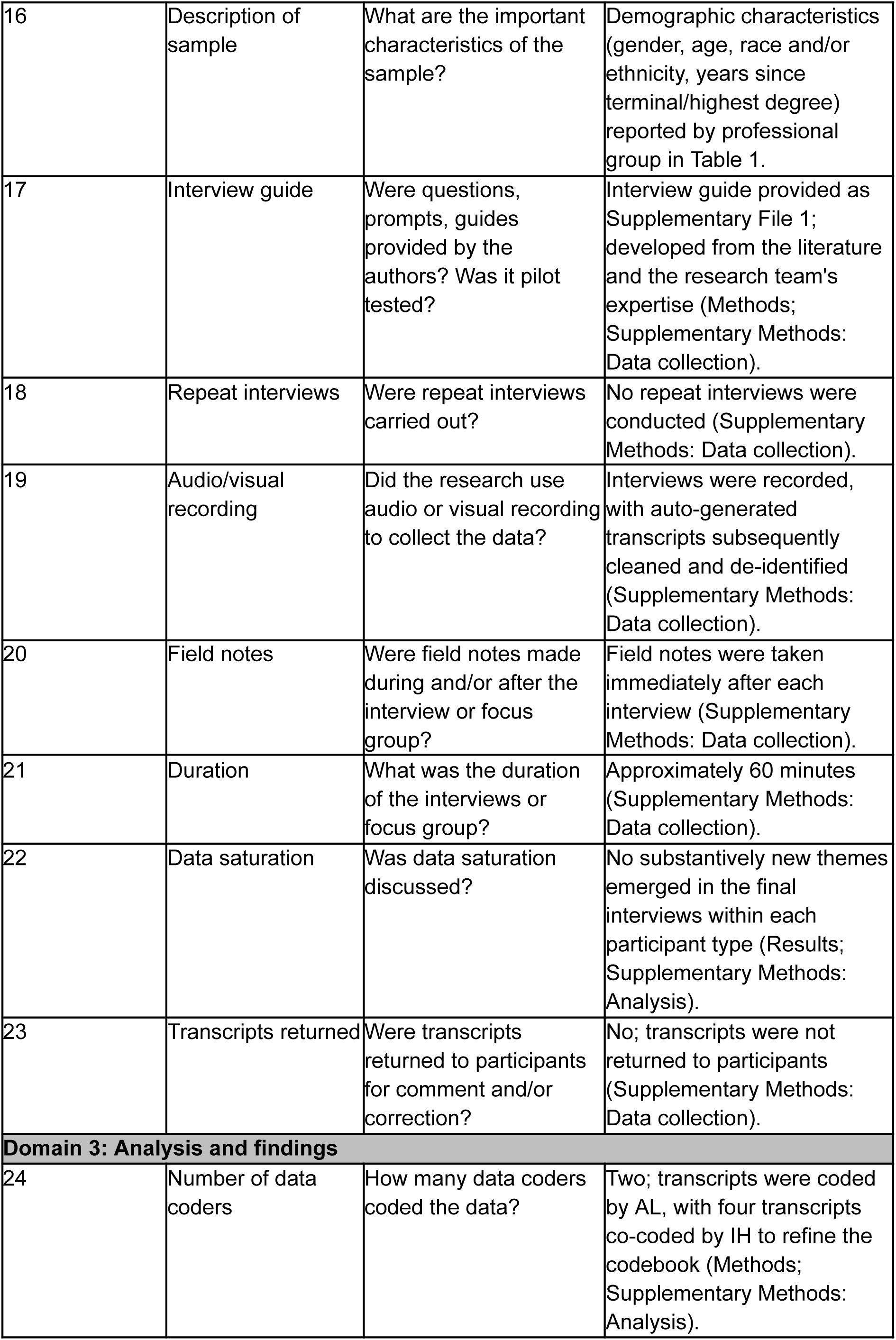
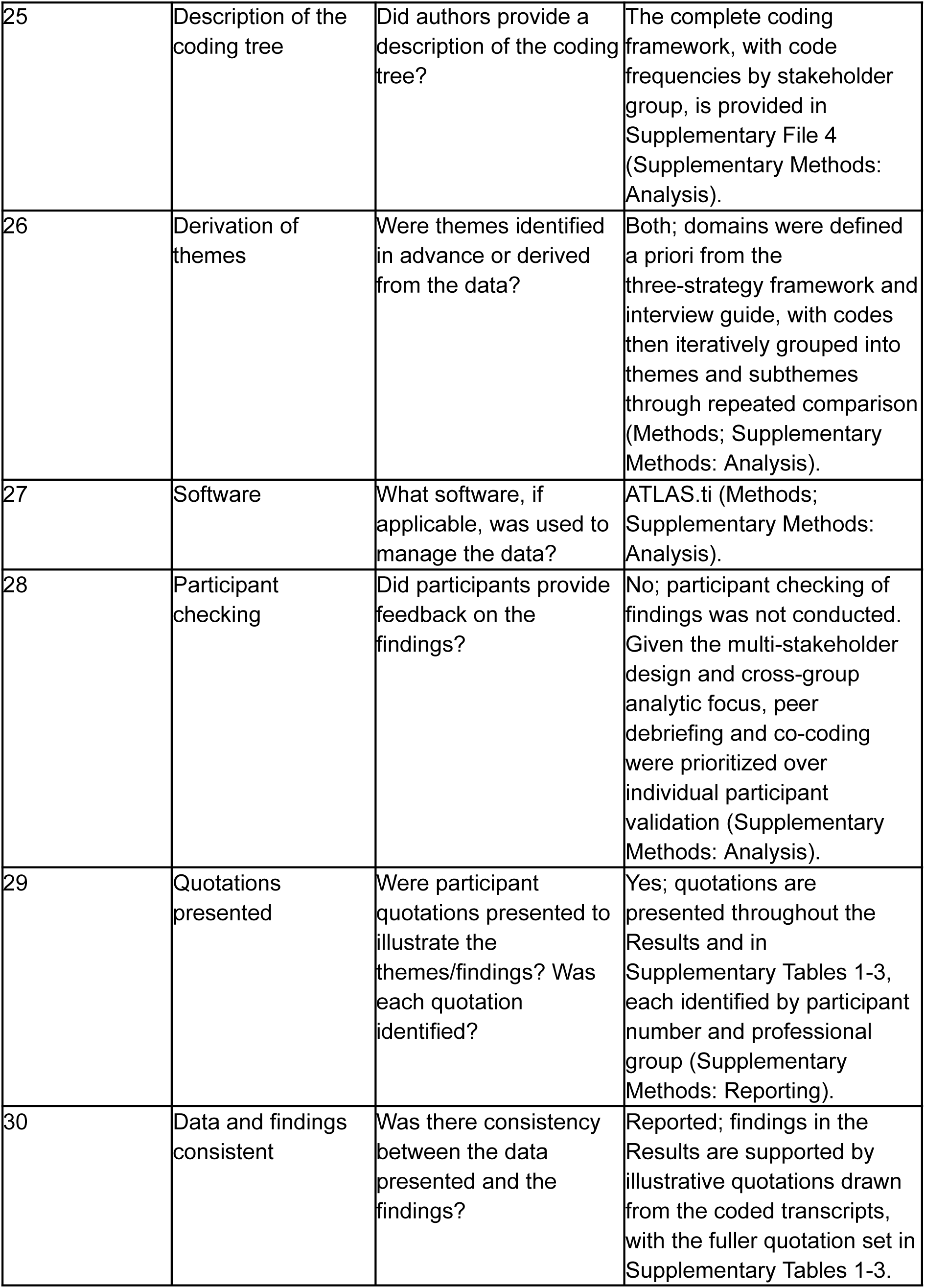
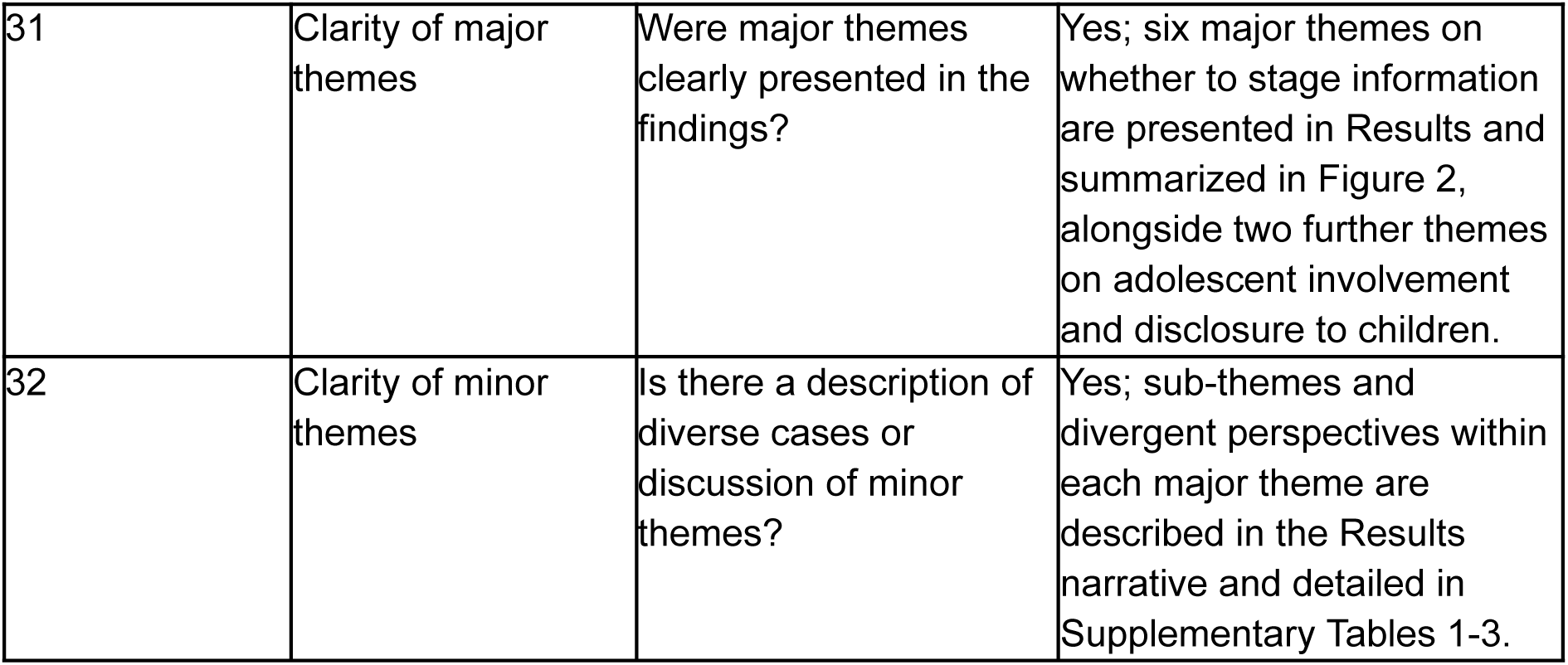

| No. | Item | Guide question/description | Reported on (section) |
| --- | --- | --- | --- |
| **Domain 1: Research team and reflexivity** |  |  |  |
| 1 | Interviewer/facilitator | Which author conducted the interview? | AL (Anna C F Lewis) conducted all interviews (Methods). |
| 2 | Credentials | What were the researcher's credentials? | AL, DPhil (Supplementary Methods: Research team and reflexivity). |
| 3 | Occupation | What was their occupation at the time of the study? | Researcher working at the intersection of genomics and ethics (Supplementary Methods). |
| 4 | Gender | Was the researcher male or female? | Female (she/her) (Supplementary Methods). |
| 5 | Experience and training | What experience or training did the researcher have? | AL and IH are described as experienced qualitative researchers (Supplementary Methods); no further training detail is specified. |
| 6 | Relationship established | Was a relationship established prior to study commencement? | A small number of participants were known to AL before the study (Supplementary Methods). |
| 7 | Participant knowledge of the interviewer | What did the participants know about the researcher? | Prospective participants received a fact sheet describing the study and its purpose before agreeing to participate (Methods; Supplementary File 2). |
| 8 | Interviewer characteristics | What characteristics were reported about the interviewer/facilitator? | AL was a partial insider to the professional community studied, which may have facilitated candor but may also have led some participants to foreground ethical considerations (Supplementary Methods: Research team and reflexivity). |
| **Domain 2: Study design** |  |  |  |
| 9 | Methodological orientation and theory | What methodological orientation was stated to underpin the study? | Framework analysis, combining deductive and inductive coding, with a pragmatist orientation (Methods; Supplementary Methods: Analysis). |
| 10 | Sampling | How were participants selected? | Purposive sampling supplemented by snowball sampling (Methods; Supplementary Methods: Sampling and recruitment). |
| 11 | Method of approach | How were participants approached? | Email invitation, individualized or via institutional listservs; laboratory directors and implementers invited directly by email (Supplementary Methods: Sampling and recruitment). |
| 12 | Sample size | How many participants were in the study? | 52 (clinical geneticists n=6; genetic counselors n=16; laboratory personnel n=8; pediatric primary care clinicians n=10; genomic screening implementers n=12) (Results; Table 1). |
| 13 | Non-participation | How many people refused to participate or dropped out? Reasons? | Partially reported: 7 of 14 invited laboratory directors participated; 12 of 18 invited implementers participated (Supplementary Methods). Response rate could not be calculated for listserv-recruited clinical groups because the number of recipients was not ascertained; no dropouts are reported. |
| 14 | Setting of data collection | Where was the data collected? | Videoconference (Zoom) (Methods; Supplementary Methods: Data collection). |
| 15 | Presence of non-participants | Was anyone else present besides the participants and researchers? | No; only the interviewer and participant were present (Supplementary Methods: Data collection). |
| 16 | Description of sample | What are the important characteristics of the sample? | Demographic characteristics (gender, age, race and/or ethnicity, years since terminal/highest degree) reported by professional group in Table 1. |
| 17 | Interview guide | Were questions, prompts, guides provided by the authors? Was it pilot tested? | Interview guide provided as Supplementary File 1; developed from the literature and the research team's expertise (Methods; Supplementary Methods: Data collection). |
| 18 | Repeat interviews | Were repeat interviews carried out? | No repeat interviews were conducted (Supplementary Methods: Data collection). |
| 19 | Audio/visual recording | Did the research use audio or visual recording to collect the data? | Interviews were recorded, with auto-generated transcripts subsequently cleaned and de-identified (Supplementary Methods: Data collection). |
| 20 | Field notes | Were field notes made during and/or after the interview or focus group? | Field notes were taken immediately after each interview (Supplementary Methods: Data collection). |
| 21 | Duration | What was the duration of the interviews or focus group? | Approximately 60 minutes (Supplementary Methods: Data collection). |
| 22 | Data saturation | Was data saturation discussed? | No substantively new themes emerged in the final interviews within each participant type (Results; Supplementary Methods: Analysis). |
| 23 | Transcripts returned | Were transcripts returned to participants for comment and/or correction? | No; transcripts were not returned to participants (Supplementary Methods: Data collection). |
| **Domain 3: Analysis and findings** |  |  |  |
| 24 | Number of data coders | How many data coders coded the data? | Two; transcripts were coded by AL, with four transcripts co-coded by IH to refine the codebook (Methods; Supplementary Methods: Analysis). |
| 25 | Description of the coding tree | Did authors provide a description of the coding tree? | The complete coding framework, with code frequencies by stakeholder group, is provided in Supplementary File 4 (Supplementary Methods: Analysis). |
| 26 | Derivation of themes | Were themes identified in advance or derived from the data? | Both; domains were defined a priori from the three-strategy framework and interview guide, with codes then iteratively grouped into themes and subthemes through repeated comparison (Methods; Supplementary Methods: Analysis). |
| 27 | Software | What software, if applicable, was used to manage the data? | ATLAS.ti (Methods; Supplementary Methods: Analysis). |
| 28 | Participant checking | Did participants provide feedback on the findings? | No; participant checking of findings was not conducted. Given the multi-stakeholder design and cross-group analytic focus, peer debriefing and co-coding were prioritized over individual participant validation (Supplementary Methods: Analysis). |
| 29 | Quotations presented | Were participant quotations presented to illustrate the themes/findings? Was each quotation identified? | Yes; quotations are presented throughout the Results and in Supplementary Tables 1-3, each identified by participant number and professional group (Supplementary Methods: Reporting). |
| 30 | Data and findings consistent | Was there consistency between the data presented and the findings? | Reported; findings in the Results are supported by illustrative quotations drawn from the coded transcripts, with the fuller quotation set in Supplementary Tables 1-3. |
| 31 | Clarity of major themes | Were major themes clearly presented in the findings? | Yes; six major themes on whether to stage information are presented in Results and summarized in Figure 2, alongside two further themes on adolescent involvement and disclosure to children. |
| 32 | Clarity of minor themes | Is there a description of diverse cases or discussion of minor themes? | Yes; sub-themes and divergent perspectives within each major theme are described in the Results narrative and detailed in Supplementary Tables 1-3. |

## Notes

### Author Declarations

The Mass General Brigham institutional review board determined the study exempt and did not require documented consent

